# K-space Optimized Spatial Temporal Architecture for Low Latency in Interventional Cardiac MRI

**DOI:** 10.64898/2026.09.21.26363257

**Authors:** Mark Buckup, Niraj Mahajan, Ana E Rodríguez-Soto, Joni Blood, Sanjeet Hegde, Eleanor L Schuchardt, Beth Printz, Hari K Narayan, Brent L Gordon, Francisco Contijoch

**Affiliations:** Department of Bioengineering, Jacobs School of Engineering, UC San Diego; Department of Computer Science, Jacobs School of Engineering, UC San Diego; Department of Radiology, School of Medicine, UC San Diego; Department of Pediatrics, School of Medicine, UC San Diego; Rady Children’s Hospital San Diego, Division of Cardiology

**Keywords:** real-time MRI, interventional cardiovascular MRI, recurrent neural network, golden-angle radial, cardiac cine

## Abstract

**Purpose:** To develop and evaluate a recurrent neural network that operates directly on undersampled golden-angle radial k-space, enabling low-latency cardiac cine reconstruction for interventional cardiac MRI (iCMR).

**Methods:** KOSTAL (K-space Optimized Spatial Temporal Architecture for Low latency) combines a per-coil k-space convolutional gated recurrent unit (convGRU), which learns a mask-conditioned view-sharing rule over the accumulated k-space, with an image-space recurrent module. It was trained and tested on the open-access multi-coil OCMR dataset using 10 simulated golden-angle spokes per frame against a 1000-spoke reference. Ablations varied recurrent module architecture and placement; temporal behavior was assessed using a imaging-plane transition test. Generalization was evaluated without retraining in 23 pediatric patients prospectively scanned during free-breathing using a radial golden-angle bSSFP sequence with binned XD-GRASP and clinical breath-held cine serving as references.

**Results:** At 10 spokes per frame, KOSTAL reached a median SSIM of 0.907 (gridding: 0.622) in under 34 ms of processing time per frame. Image quality (IQ) matched gridded results formed from 7-11 times more spokes. Recurrence in k-space rather than image space accounted for most of this gain (0.891 versus ≤0.650). IQ recovered from an abrupt plane transition in 75 ms, versus 303 ms for 200-spoke gridding. KOSTAL had the best agreement with the binned reference and the highest long-axis CNR. KOSTAL accurately tracked left-ventricular blood-pool volume (end-diastolic volume bias-0.7 mL) with a shorter lag (71 ms) than iterative reconstruction (239-287 ms).

**Conclusion:** Learned, mask-conditioned recurrence in k-space yields high image quality at a latency compatible with interventional guidance.

## Introduction

Accurate depiction of cardiac motion from single-shot CMR data is necessary in applications such as interventional CMR (iCMR). As the heart is constantly moving, a high number of k-space samples used for reconstruction results in cardiac blurring^1^. Therefore, image quality is often limited in single-shot CMR as reconstruction requires processing of highly undersampled k-space^2^. Techniques such as parallel imaging, iterative reconstruction, and compressed sensing/sparsity have successfully improved single-shot CMR image quality by reducing the number of views needed^3–9^. However, long reconstruction times preclude use of certain algorithms in real-time display scenarios such as iCMR, where low latency (<200 ms) is needed^5,10–12^.

Neural networks offer a potential solution: training frontloads most of the computation so inference of a trained network can be very quick on current hardware^13–17^. Further, during real-time imaging, a time series, or movie, with high temporal similarity between successive frames is generated. Neural networks can exploit this similarity to reconstruct high-quality images despite high undersampling. Recent work by El-Rewaidy et al. proposed a Multi-Domain Convolutional Neural Network (MDCNN) to reconstruct high-quality images from highly undersampled radial k-space^18^. While the MDCNN approach has several advantages, it was not specifically designed for real-time display in iCMR. For example, as the sliding window of the MDCNN shifts across the time series, each frame undergoes multiple forward passes, which increases the computational burden. In addition, the temporal window evaluated by the network was a user-defined parameter set during training and fixed afterwards. While this helps standardize the extent of undersampling, it makes the approach susceptible to variations in heart rate across patients^11,19^.

Recurrent neural networks (RNNs) are designed to model temporal patterns, so they are better suited for inference on time series^20,21^. However, they can be more difficult to train, as RNNs can suffer from vanishing gradients. This was previously addressed by “long short-term memory” (LSTM) networks, which were initially proposed for 1D time series and have since been extended to convolutional LSTMs (convLSTMs) for use on 2D videos (time series of 2D frames)^22–24^. Gated recurrent units (GRUs) provide a streamlined gated formulation with fewer parameters and comparable temporal modeling^25^. For spatiotemporal data, a convolutional form (convGRU) has been developed^26,27^. However, to date, these recurrent architectures have largely been developed for image-domain inputs. Therefore, their application to k-space data has not been reported.

Applying a recurrent network directly to k-space samples is particularly attractive because it allows the network to learn how to combine newly acquired samples with retained samples from prior frames. This builds on long-standing concepts in dynamic MRI^21,28^. However, view-sharing and keyhole methods perform this combination using fixed, hand-designed operations^29–32^. A recurrent unit operating on k-space can learn a “smart” combination operation that leverages information about which locations were actually measured in the current frame, with the goal of optimizing image quality.

In this paper, we develop and evaluate KOSTAL (K-space Optimized Spatial Temporal Architecture for Low latency), a dual-domain recurrent network built on convGRUs. At each frame, KOSTAL merges the small set of newly acquired radial spokes (10 spokes) with a recurrent state (accumulated k-space from prior frames). Importantly, the locations of the new spokes are provided alongside the data as a binary sampling mask, allowing the k-space module to distinguish measured from missing locations. In this case, the extent of prior data contributing to the reconstruction is implicitly set by the learned gating rather than by a user-defined window, and can vary across k-space with the recency of sampling at each location. The k-space module densifies k-space per-coil before a Fast Fourier Transform (FFT) is used to create an image estimate. The image estimate is then refined using a second convGRU module operating in the image domain. Supervising both intermediate representations enhances explainability. Importantly, KOSTAL operates independently across coils, enabling flexibility in the number of coils processed, adaptability to patients of varying sizes, and compatibility with other current state-of-the-art coil-combination methods.

We developed and trained the network on the open-access multi-coil OCMR dataset^33^, in which golden-angle radial sampling was retrospectively simulated. Once trained, we demonstrated its clinical utility by applying the network to prospectively acquired free-breathing golden-angle radial acquisitions in a pediatric cohort. Reconstruction accuracy was quantified against fully sampled gridded references in the simulated studies and against state-of-the-art reconstructions in the in vivo studies.

## Methods

### KOSTAL Overview and Design

KOSTAL is a dual-domain, recurrent network that reconstructs cardiac cine MR images from undersampled multi-coil radial k-space (Figure 1A). Multi-coil cardiac data are processed coil-wise in k-space and image-space before coil combination. In training, testing, and validation, virtual coils were provided as inputs. Complex k-space samples are represented as separate log-magnitude (base 10) and phase inputs processed by independent recurrent pathways, so amplitude and phase are recovered in the acquired domain before image formation. The binary sampling mask is input alongside the undersampled data throughout, making the network explicitly aware of which locations were measured.

**FIGURE 1:**
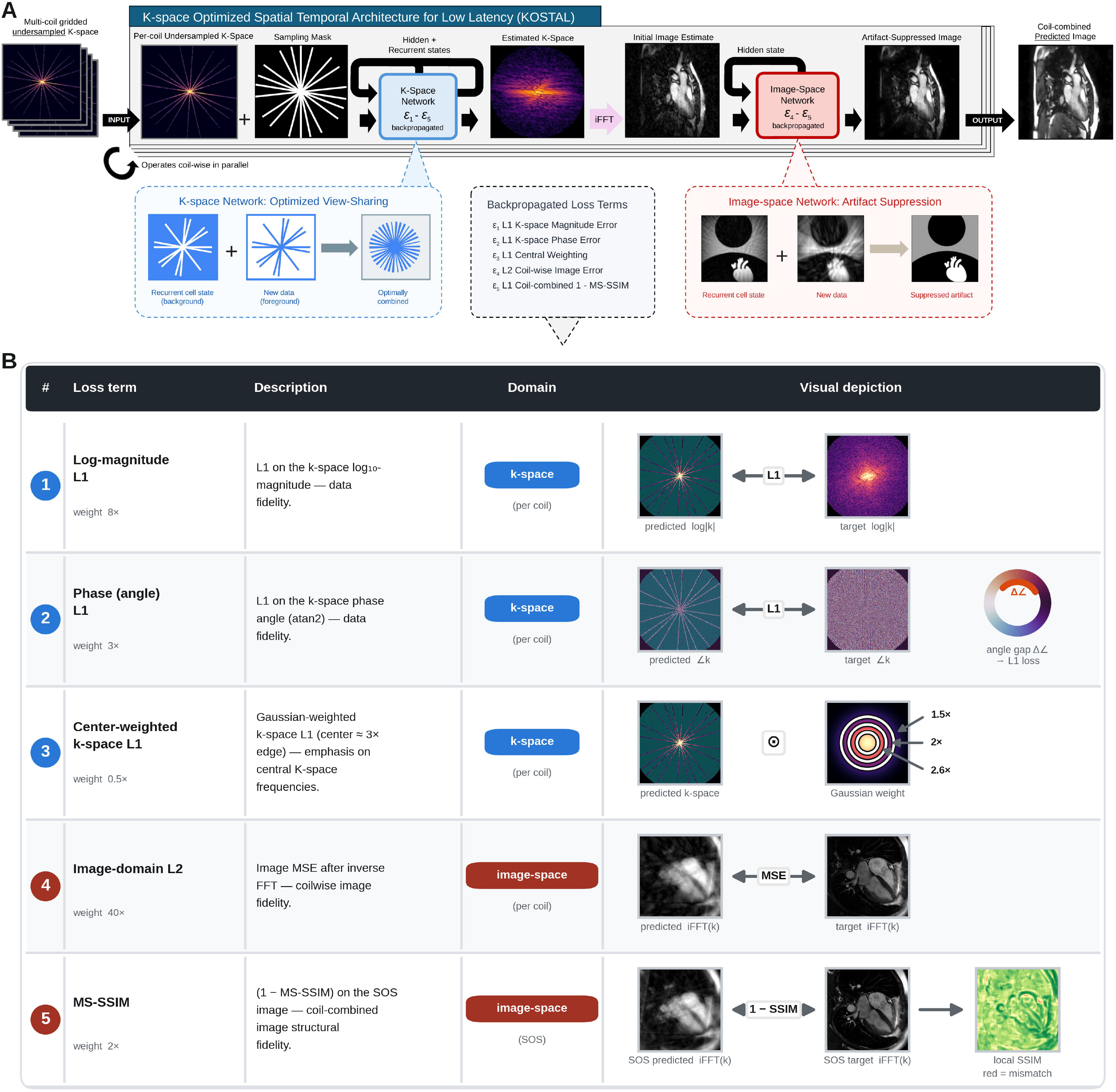
KOSTAL architecture and training objective. **(A)** Per-coil gridded radial k-space and its sampling mask enter a recurrent k-space network, which merges the state carried forward from preceding frames with the spokes newly acquired for the current frame (left) and returns a densified k-space estimate. After inverse Fourier transform (iFFT), a second recurrent network suppresses the residual undersampling artifact (right), and the per-coil images are combined into the predicted image. Both networks carry a hidden state between frames; all five loss terms are backpropagated into the k-space network, and the two image-domain terms are additionally backpropagated into the image-space network. **(B)** The five loss terms expanded from the dashed box of (A): weight, description, domain (k-space, blue; image space, red), granularity (per-coil or SOS combination), and a visual example on multicoil data.

The network comprises two learned recurrent stages. First, a per-coil k-space convolutional gated recurrent unit (convGRU; 4 convolutional layers, 64 hidden channels, unbounded temporal context) integrates the current acquisition into a k-space state accumulated from prior radial frames, densifying the k-space of each virtual coil. A gate-consistency term constrains the candidate state and the gated output to agree with the measured samples, thereby preserving the new data when combined with the prior memory. Therefore, in this step, the network learns how to incorporate consistent background context. Coil-resolved k-space estimates are then inverse Fast Fourier transformed (iFFT). Then, in the image-domain, a per-coil image-space convGRU initialized using the identity operation (48 hidden channels, 2 convolutional layers per gate) refines the magnitude images. The final multi-coil image stack is generated, defaulting to a standard sum-of-squares (SOS) coil combination for display.

### Data Acquisition and Preparation for Training

Model training and testing was performed using the Open-access Multi-Coil k-space (OCMR) dataset from Ohio State University. OCMR provides true multi-coil k-space acquired on Siemens MAGNETOM scanners at three field strengths (0.55 T Free.Max, 1.5 T Avanto/Sola, and 3 T Prisma) using breath-held, ECG-gated bSSFP cine imaging. Fully sampled cine series (n=165 series and n=279 imaging slices) spanning short-axis (SAX) and long-axis (LAX) views were used.

Golden-angle radial sampling was retrospectively simulated. Readout oversampling was removed using the header matrix size and field of view, and, where noise scans were available, coils were prewhitened with an estimated noise-decorrelation matrix. Receiver channels were compressed to 8 virtual coils by singular-value decomposition (SVD)^34,35^. This standardizes the network input and was required for comparison with the MDCNN and Gadgetron-based architectures. Each slice contained 10 to 31 acquired cardiac phases (mean 22). For simulation of real-time single-shot imaging, series were cyclically repeated to form 120-frame sequences.

Golden-angle radial sampling was simulated with a random start index per slice, and 10 spokes were sampled from each ECG-gated cardiac phase-resolved k-space with per-frame intensity normalization. For comparison, a dense 1000-spoke acquisition per cardiac phase was used as the fully sampled radial reference. Undersampled and reference radial k-space data were density-compensated and re-gridded onto a 256×256 Cartesian matrix by Kaiser-Bessel adjoint NUFFT interpolation on a 4x oversampled grid, then decimated. Coil-wise undersampled and reference k-space, the normalized SOS target image, and a binary spoke mask were saved per frame.

### Network Training

Datasets were partitioned 80/20 into training and testing sets, yielding 132 training and 33 testing patients (237 and 42 imaging slices, all slices from a patient were kept within their respective splits). Training used clips of 20 consecutive frames, with random starting frames, for temporal augmentation; the first 8 frames were used for recurrent warm-up and were excluded from the loss. The model was trained for 200 epochs (batch size 1, Adam, betas 0.9 and 0.999) in two phases: (1) the k-space GRU was trained alone for the first 100 epochs, (2) then the image-space GRU was activated and the two were trained jointly through epoch 200. Both used an exponential-range cyclic learning rate (base 4e-6, maximum 6e-4, 2000 steps to maximum, decay 0.99995) with gradient norms clipped at 0.8. No early stopping was used.

Supervision was performed using multiple terms across both domains. The k-space module backpropagated five losses: a) coil-wise k-space log-magnitude L1; b) coil-wise k-space phase angle L1; c) coil-wise Gaussian center-weighted k-space L1^36^ (weighting the k-space center approximately threefold relative to the edge); d) a coil-wise image-domain L2; and e) multiscale structural similarity index measure (MS-SSIM) loss on the SOS of the k-space output (11×11 spatial window). The image-space module backpropagated the latter two loss terms (d and e). Relative weights are given in Figure 1B and were chosen to balance coil-wise and coil-combined quality against recurrent temporal behavior and recovery speed (Figure S1).

### Training and Testing Evaluation Criteria

Reconstructions were evaluated against the 1000-spoke fully sampled radial reference on the 33 held-out test series using complementary similarity and error metrics (Figure 2A-B). Structural similarity index (SSIM) was computed with an 11×11 local window to quantify local structural agreement. Mean-squared error (MSE) was derived from the mean squared intensity difference to give an absolute pixel-wise error. To evaluate the performance of each component, metrics were computed after application of three different forward-pass variants: (1) a k-space-only processing output (iFFT of the k-space-GRU output), (2) an image-space-only processing output (the image-space GRU was applied directly to the iFFT of the undersampled input, bypassing the k-space module), and (3) the combined k-space-plus-image-space output. Coil-wise metrics assessed the per-coil intermediate reconstructions, while SOS metrics assessed the final coil-combined image. Analyses were performed on both the full image and a central 50% region of interest (ROI) focused on the heart.

**FIGURE 2:**
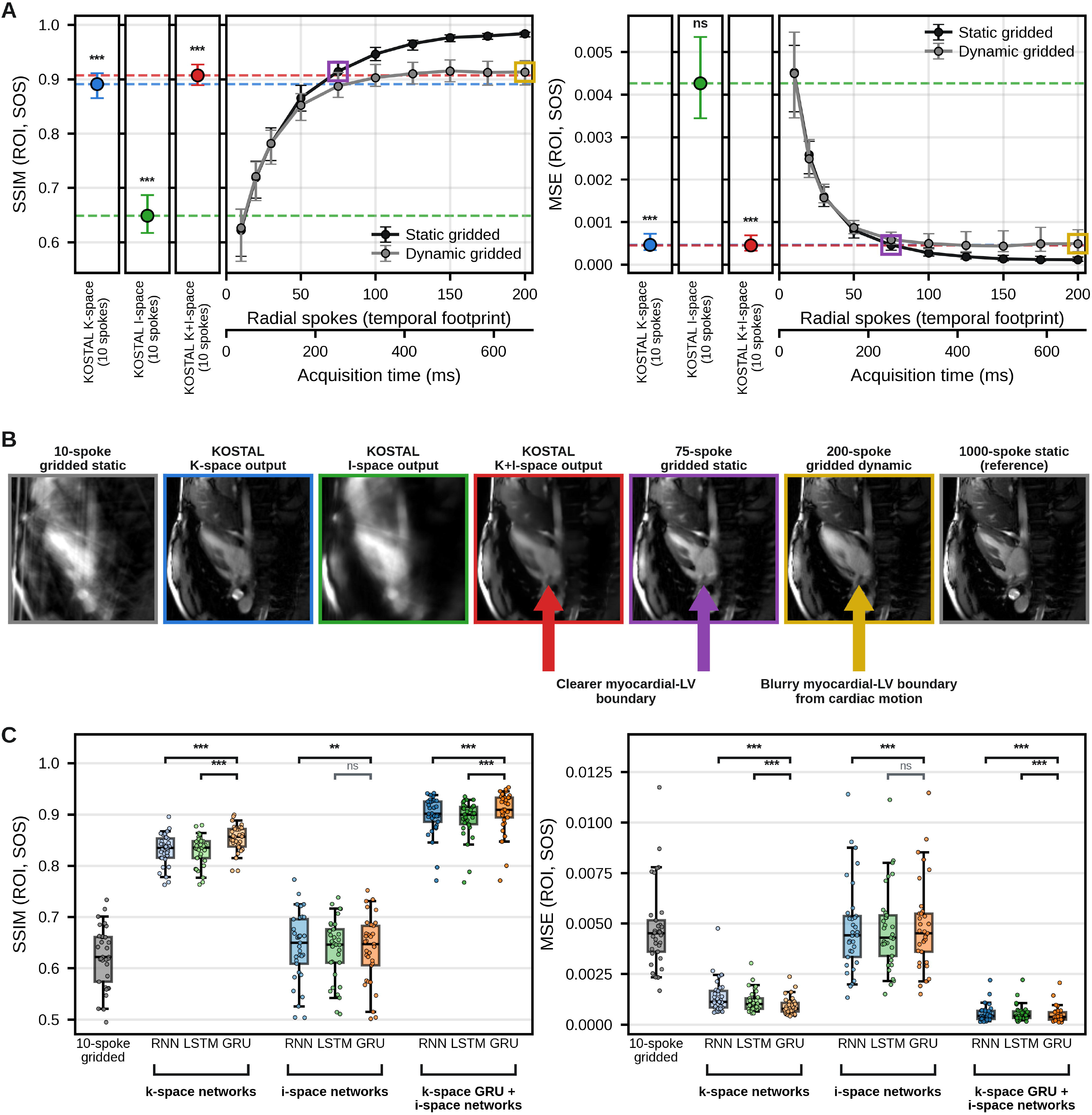
Reconstruction accuracy as a function of sampling density in a simulated retrospective cohort. **(A)** Structural similarity index (SSIM, left) and mean squared error (MSE, right) over the central 50% ROI of the SOS combination against the number of radial spokes gridded per frame. Black, static gridding, in which every spoke of a frame comes from one retrospectively binned reference frame (theoretical best case); gray, dynamic gridding, in which spokes accrue along the golden-angle trajectory (real-time case, in which a longer temporal footprint additionally blurs cardiac motion). Symbols are medians, and error bars are the first to third quartiles. The three narrow columns at left give the k-space, image-space, and combined KOSTAL outputs from a 10-spoke input, and the dashed horizontal lines carry those medians across the gridded curves so the equivalent spoke count can be read off directly. The lower axis converts spoke count to acquisition time. The open purple and gold squares mark the two gridded operating points imaged in (B). **(B)** One cardiac frame of a representative held-out patient reconstructed seven ways, with border colors as in (A); the three arrows mark the same myocardium-to-blood-pool boundary. **(C)** Ablation of the recurrent cell (plain RNN, LSTM, GRU). Boxes span the first to third quartiles, with the median marked, whiskers at the 5th and 95^th^ percentiles, and each dot represents one patient. Asterisks in (A) give the significance of each network output against the 10-spoke gridded input, and brackets in (C) compare the gated recurrent unit with the other two cells within each group, both by a two-sided Wilcoxon signed-rank test over the same 33 patients with Holm correction within the comparison family: *** p<0.001, ** p<0.01, * p<0.05, ns not significant.

Two different gridded radial reconstructions, performed with different numbers of radial spokes, yielded reference error values. In the static gridded reconstruction, N golden-angle spokes of a frame came from a *single* retrospectively binned cardiac phase. Therefore, increasing N adds spoke density without motion blur. This represents the theoretical best-case scenario for a given spoke budget but is not achievable in practice. In the dynamic gridded reconstruction, spokes are accrued as in a *continuous* real-time acquisition. Therefore, larger N lengthens the temporal footprint, leading to incorporation of spokes from different cardiac phases. This represents conventional reconstruction of different window widths. Both were evaluated from 10 to 1000 spokes. The gridded-spoke equivalent in Table 1 is the largest spoke count, up to the 1000-spoke ceiling, at which the gridded values match the proposed method (linearly interpolated between tested spoke values).

**TABLE 1:** Reconstruction image quality in the ROI for SOS and coil-wise average coil-combination strategies. Values are the per-patient median (first to third quartile) over n = 33 held-out test patients. Metrics were computed inside the central 50% ROI, which contains the heart, relative to the fully sampled reference; SSIM and MSE are dimensionless, and MSE is reported multiplied by 10-3. Gridded-spoke equivalent is the largest number of radial spokes, up to the 1000-spoke ceiling of the acquired data, at which a gridded reconstruction reached the same median value, interpolated linearly between sampled spoke counts. “Static” denotes gridding from a static acquisition model and “dynamic” gridding in real time. Asterisks give the significance of each KOSTAL output against the 10-spoke gridded reconstruction of the same block, by a two-sided Wilcoxon signed-rank test over the same 33 patients with Holm correction within each metric and coil-combination strategy: *** p<0.001, ** p<0.01, * p<0.05, ns not significant.

| Metric | Reconstruction | Sum-of-squares: median<br>(Q1–Q3) | Sum-of-squares: gridded-spoke equivalent, static | Sum-of-squares: gridded-spoke equivalent, dynamic | Coil-wise average: median<br>(Q1–Q3) | Coil-wise average: gridded-spoke equivalent, static | Coil-wise average: gridded-spoke equivalent, dynamic |
| --- | --- | --- | --- | --- | --- | --- | --- |
| *SSIM* | Gridded, 10 spokes | 0.622 (0.574–0.661) | 10 | <10 | 0.780 (0.742–0.800) | 10 | 11 |
| *SSIM* | KOSTAL, k-space output | 0.891 (0.866–0.911) | 63 | 81 | 0.921 (0.876–0.943) | 49 | 64 |
| *SSIM* | KOSTAL, image-space output | 0.649 (0.617–0.687) | 13 | 12 | 0.803 (0.765–0.829) | 14 | 15 |
| *SSIM* | KOSTAL, combined output | 0.907 (0.889–0.927) | 71 | 114 | 0.945 (0.915–0.960) | 72 | 211 |
| *MSE* ($\times 10^{-3}$) | Gridded, 10 spokes | 4.524 (3.604–5.157) | 10 | <10 | 0.658 (0.515–0.846) | 10 | <10 |
| *MSE* ($\times 10^{-3}$) | KOSTAL, k-space output | 0.467 (0.339–0.723) | 74 | 223 | 0.106 (0.064–0.168) | 64 | 78 |
| *MSE* ($\times 10^{-3}$) | KOSTAL, image-space output | 4.269 (3.448–5.356) | 11 | 11 | 0.597 (0.449–0.765) | 12 | 12 |
| *MSE* ($\times 10^{-3}$) | KOSTAL, combined output | 0.454 (0.325–0.687) | 76 | 235 | 0.081 (0.055–0.142) | 76 | 412 |

To test the appropriateness of the proposed architecture, two ablations were performed. First, the recurrent cell was varied between a plain convRNN, a convLSTM cell, and the proposed convGRU, in three configurations: (1) the k-space module alone, (2) an image-space module applied directly to the gridded input, and (3) the dual-domain arrangement (Figure 2C). Second, performance after ablation of each of the five loss terms of Figure 1B are reported (Figure S1).

Temporal behavior was assessed using an abrupt change in imaging plane: 33 held-out LAX-to-SAX transition pairs were constructed and scored frame-by-frame against the 1000-spoke reference across the transition (Figure 3). Recovery time was defined per pair as the time after the plane change at which a reconstruction reached the steady-state quality achieved by 10-spoke gridding. This was calculated as the mean of the last 10 frames of that pair’s 10-spoke gridded curve. The performance of KOSTAL was compared with gridded reconstructions at 10-, 75-, and 200-spoke temporal footprints under both SSIM and MSE criteria. One frame corresponds to 34 ms, since the TR was simulated as 3.35 ms for each spoke.

**FIGURE 3:**
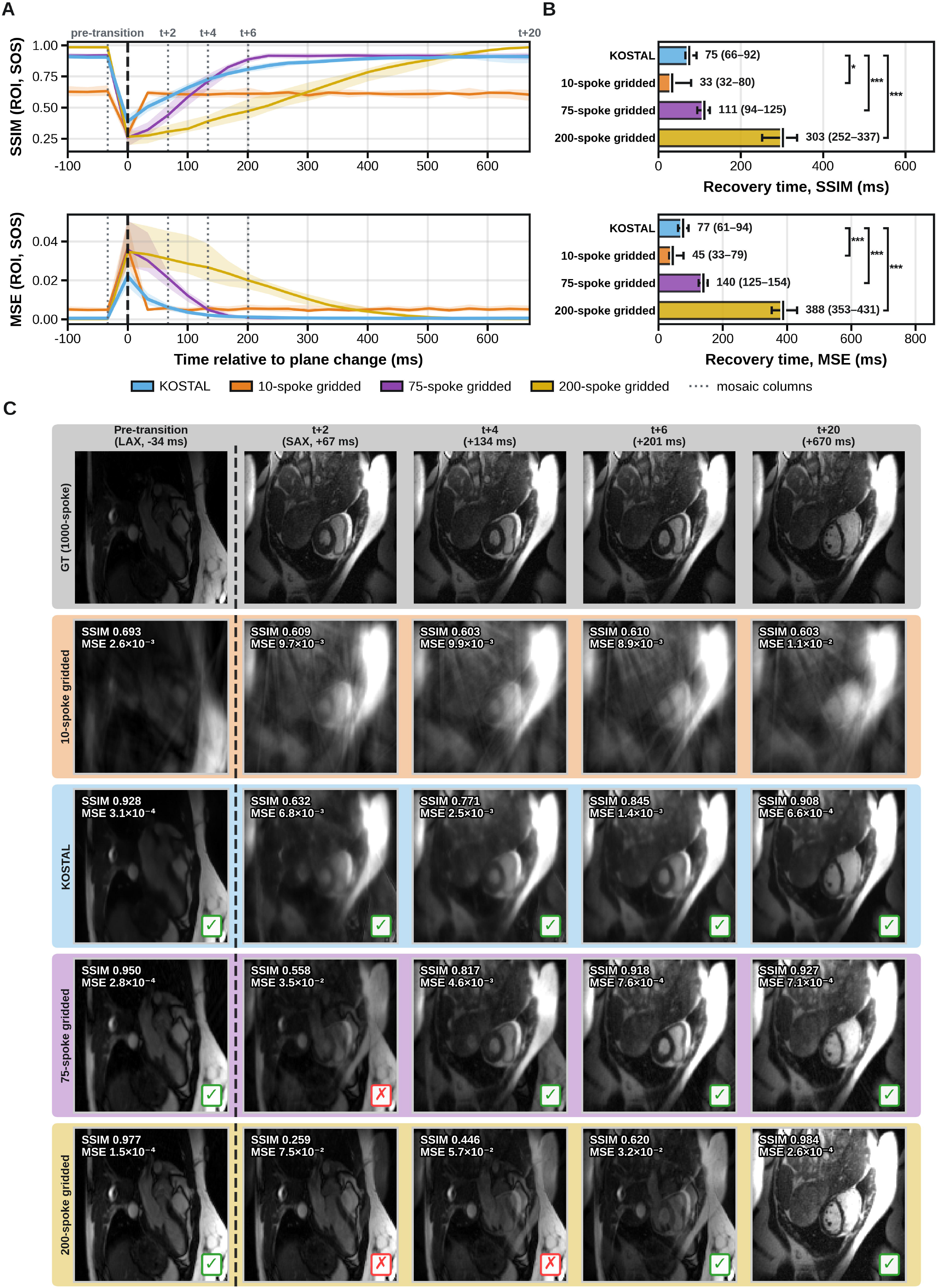
Behavior of each reconstruction across an abrupt change of imaging plane. (A) Median SSIM (top) and MSE (bottom) over the central 50% ROI of the SOS combination, against time relative to the plane change at 0 ms (dashed vertical line); bands include 25th to 75th percentiles and dotted vertical lines mark the five time points imaged in (C). One frame is 34 ms. (B) Recovery time, defined per pair as the first time after the plane change at which a reconstruction reached the steady-state quality that 10-spoke gridding eventually attains for that pair, by the SSIM (top) and MSE (bottom) criteria. Bars are medians, and whiskers are the first to third quartiles. Because the target is the 10-spoke gridded steady state itself, that bar is a self-reference and is shown only for completeness. (C) One representative LAX-to-SAX transition at the five marked time points, one row per reconstruction, with the dashed line separating the pre-and post-transition columns. The mark in the lower right of each tile indicates whether that reconstruction met the SSIM of 10-spoke gridding in the same column; no mark is placed on the reference or comparator rows. Brackets to the right of the bars compare the proposed network with each gridded footprint by a two-sided Wilcoxon signed-rank test over the same 33 transition pairs, Holm-corrected within each row: *** p<0.001, ** p<0.01, * p<0.05, ns not significant.

### Validation Dataset and Preprocessing

Generalization was assessed in an independent cohort of N=23 pediatric patients scanned at Rady Children’s Hospital San Diego (RCHSD) under an institutional review board-approved protocol, with written informed consent and assent. Patients undergoing clinically indicated cardiac MRI without anesthesia were approached consecutively for additional research imaging during the same session. Each underwent free-breathing 2D golden-angle radial cine imaging: a multi-slice SAX stack and, for most (N=20), single-slice LAX two-, three-, and four-chamber scans. Demographics, diagnoses, and acquisition parameters are given in Table 2. Unlike OCMR, no fully sampled reference exists for these acquisitions.

**TABLE 2:**
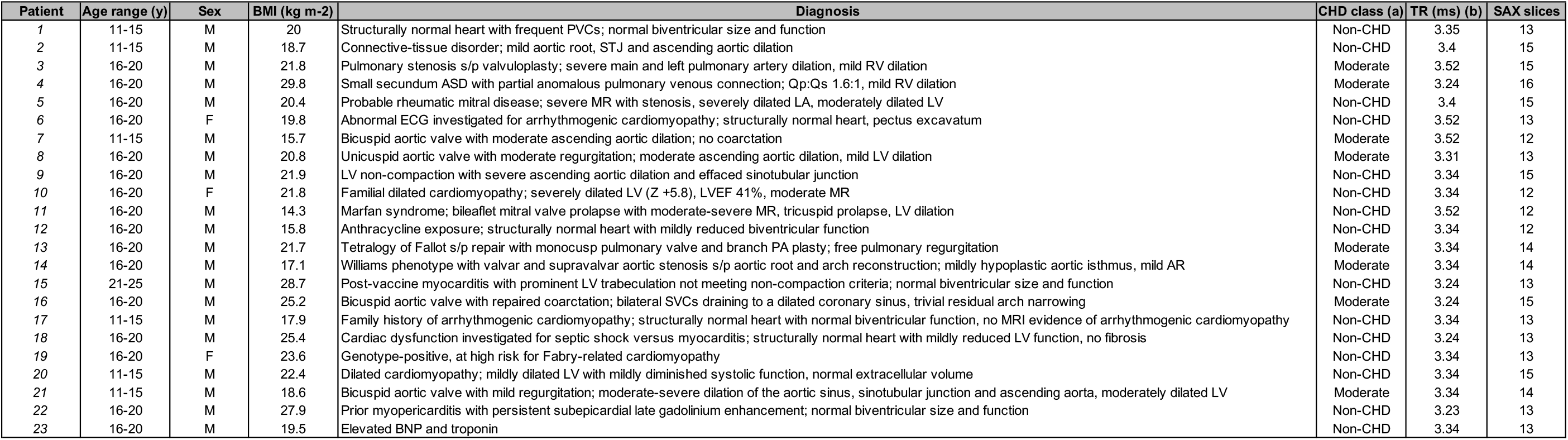
Prospective validation cohort: demographics, diagnosis and acquisition parameters. ^a^. CHD class is the anatomic axis of the AHA/ACC anatomic and physiological classification of congenital heart disease, which grades congenital anatomy as simple, moderate complexity, or great complexity. The axis ranks congenital anatomy only, so acquired, genetic-aortopathic, and cardiomyopathic disease lies outside it and is reported as Non-CHD. ^b^TR is the per-readout repetition time taken from the ISMRMRD echo-spacing field; TE was TR/2 in every scan, as expected for bSSFP. Abbreviations: 2CH, two-chamber; 3CH, three-chamber; 4CH, four-chamber; BMI, body mass index; CHD, congenital heart disease; NA, not available; SAX, SAX.

Preprocessing used the same 256×256 NUFFT gridding, with channels compressed to 8 virtual coils using region-optimized virtual (ROVir) coil compression (1000 calibration spokes, regularization 0.5, circular target region of radius one-quarter of the matrix size)^37^. While training employed SVD-based coil compression, validation used ROVir to better preserve signal energy and introduce variation between the training and validation datasets, thereby improving KOSTAL’s robustness (Figure S2). Each acquisition was reconstructed with KOSTAL and compared against a multidomain convolutional neural network (MDCNN), Gadgetron locally low-rank plus sparsity (LALM) reconstructions at 10-and 32-spoke (default) temporal footprints, and gridded radial baselines (Figure 4A-B). Both LALM variants were run causally, using only already-acquired spokes. An internal multi-shot, respiratory-and cardiac-phase binned reference standard was formed per acquisition using the XD-GRASP pipeline (20 cardiac by 2 respiratory bins)^38^. Clinical breath-held, retrospectively gated bSSFP SAX cine served as an external reference for volumetric comparison.

**FIGURE 4:**
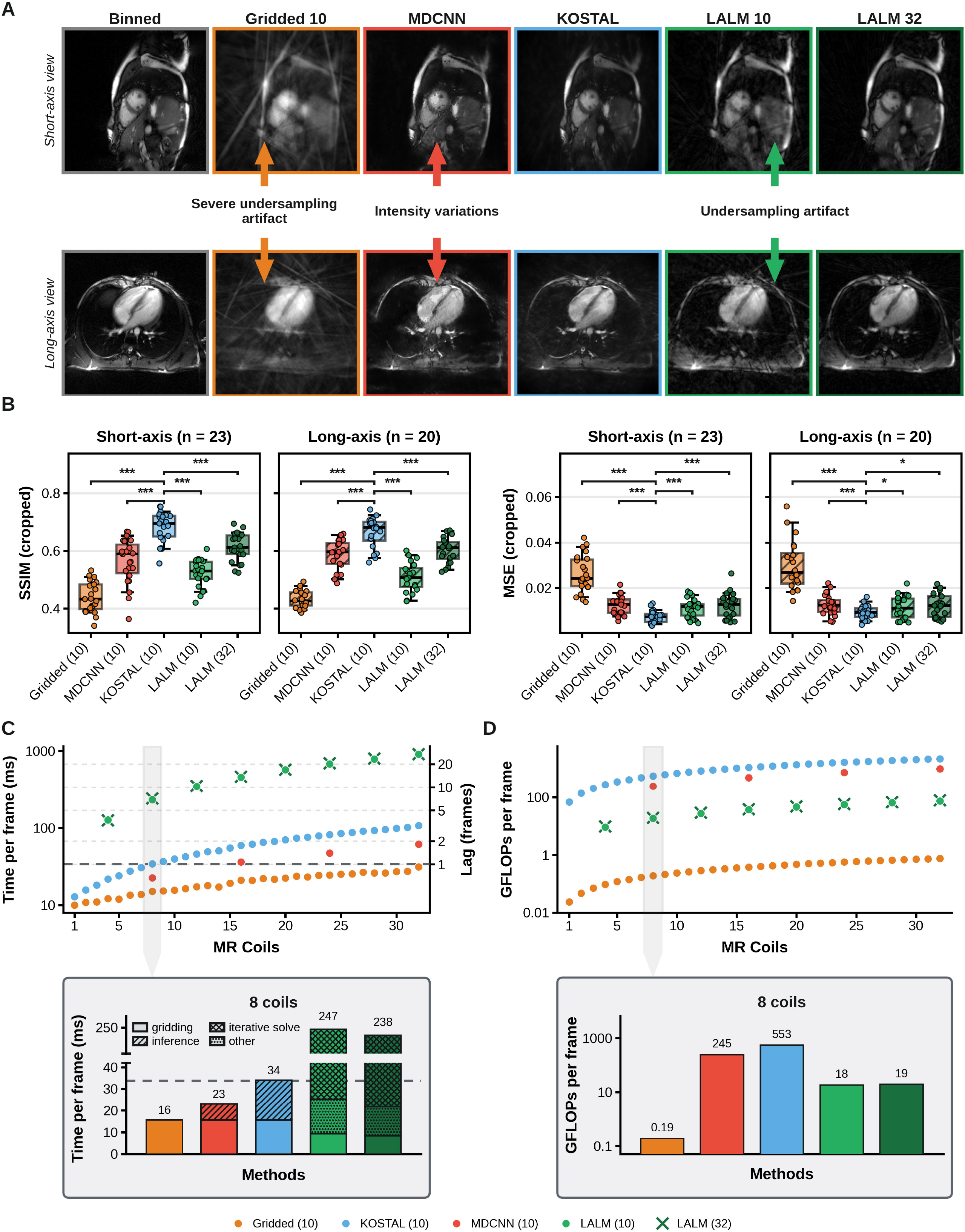
Prospective in vivo image quality and computational cost. **(A)** The same cardiac frame reconstructed by every method, for one SAX acquisition (top row) and, in a second patient, one four-chamber LAX acquisition (bottom row), alongside the respiratory-and cardiac-binned reference (Binned). Methods are 10-spoke gridding (Gridded 10), MDCNN, KOSTAL, and Gadgetron locally low-rank plus sparsity iterative reconstruction (LALM) at 10-and 32-spoke temporal footprints. Arrows mark the residual streaking of 10-spoke gridding, the intensity variation left by MDCNN, and the undersampling artifact remaining in the 10-spoke iterative reconstruction. **(B)** SSIM and MSE against the binned reference over a cropped cardiac ROI. Each point is one patient, taken as the median over that patient’s slices and frames; SAX solid boxes (n = 23), LAX hatched boxes (n = 20, pooling the two-, three-, and four-chamber views). Boxes span the first to third quartiles, with the median marked and whiskers extending to the 5th and 95th percentiles. **(C)** Total reconstruction time per frame, including the gridding step common to every method; error bars are one SD. **(D)** Inference cost per frame in giga floating-point operations (GFLOPs) on a logarithmic axis; operation counts are deterministic for a fixed input size and carry no variance. (C) and (D) exclude the once-per-acquisition noise prewhitening and coil compression. Brackets compare the proposed network with each other reconstruction by a two-sided Wilcoxon signed-rank test over the patients contributing to that axis, Holm-corrected within each axis; no comparisons were made among the other reconstructions. *** p<0.001, ** p<0.01, * p<0.05, ns not significant.

### Validation using Image Quality

Computational cost and time were measured during inference for 256×256 pixels and 10 spokes per frame across different numbers of processed coils (Figure 4C-D). Floating-point operation counts (FLOPs) were derived from operator-level tracing and CUDA event timings. Gridding time was measured with density-compensation weights computed once and reused. The iterative reconstructions were timed inside the Gadgetron gadget. Timing values exclude the once-per-acquisition pre-whitening and coil compression. All computed values are reported per coil-combined frame. All timings were performed on a Ryzen Threadripper PRO 9975WX workstation with 32 cores (64 threads), 384 GB of RAM, and 2 NVIDIA RTX PRO 6000 Blackwell Max-Q Workstation Edition GPUs (96 GB each). MDCNN (pretrained for n=8) and LALM (n=4 to reduce latency) required a specific multiple of coils, whereas gridded and KOSTAL could accommodate any number of coils.

SSIM and MSE against the binned XD-GRASP-derived reference were computed over the cardiac ROI. Contrast-to-noise ratio (CNR) and edge sharpness were estimated using ROIs manually drawn for each acquisition, specifically containing the heart (Figure 5). Specifically, a blood-pool signal region and a line profile crossing the myocardial border were manually drawn. Contrast-to-noise ratio (CNR) was defined as the difference between the maximum and minimum pixel value along the profile divided by the standard deviation within the signal region. Edge sharpness (ES) was defined as the maximum intensity gradient along the same profile. Both are reported as percentages of the same patient’s binned reference, allowing patients with different baseline contrast levels to be pooled. The median over that patient’s slices and frames was calculated with SAX and LAX acquisitions reported separately.

**FIGURE 5:**
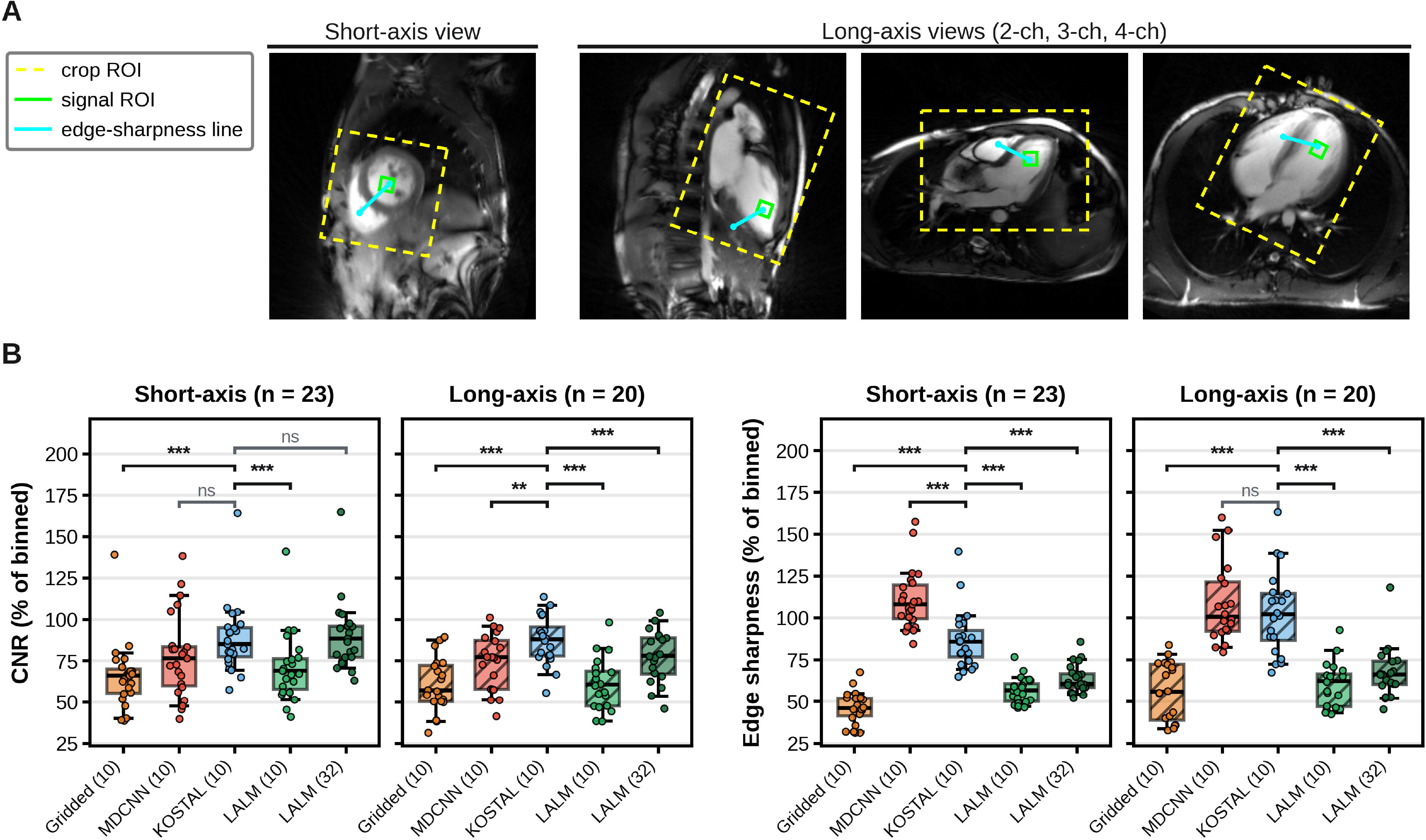
Analysis regions and reference-free image quality in the prospective validation cohort. **(A)** Regions used for the quantitative analysis, drawn on a binned reference frame from each acquired view (SAX, left; two-, three-, and four-chamber LAX, right), each tile from a different patient. Dashed box, crop over which the cardiac-focused full-reference metrics of Figure 4B were computed; solid box, blood-pool signal region from which contrast-to-noise ratio (CNR) was derived; line, myocardial profile along which edge sharpness was measured. **(B)** CNR (left) and edge sharpness (right) for each reconstruction, expressed as a percentage of the same patient’s binned reference. Each point represents one patient, taken as the median across that patient’s slices and frames. Boxes span the first to third quartiles, with the median marked and whiskers extending to the 5th and 95th percentiles. Brackets compare the proposed network with each other reconstruction by a two-sided Wilcoxon signed-rank test over the patients contributing to that axis, Holm-corrected within each axis; no comparisons were made among the other reconstructions. *** p<0.001, ** p<0.01, * p<0.05, ns not significant.

### Validation using Volume over Time Curves

Following video reconstruction, each method’s latency was explicitly calculated. Specifically, the time series was shifted by its reconstruction latency to reflect the delay an operator would experience in real time. Because the binned reference is non-causal, no delay was applied (Video S1).

Left-ventricular (LV) slice area (and measures of function) was then quantified by automated slice-volume segmentation for all methods (Figure 6, Figure S3). The LV blood pool was labeled throughout the cardiac cycle using a frozen MedSAM2 video-segmentation model (SAM2.1 Hiera-tiny backbone), with parameters manually optimized to segment the LV cavity across 36 OCMR slices (mean Dice = 0.91)^39^. For each validation patient slice, the LV centroid was manually identified and used as the segmentation seed across the time series for MedSAM2. Per-slice LV volume was computed as the segmented cavity area multiplied by the in-plane pixel area and slice thickness, reported over time for one cardiac cycle, with end-diastolic, mid-systolic, and end-systolic events taken from the volume-time curve. The mean TR over the 23 SAX acquisitions was 3.3725 ms, yielding a frame period of 34 ms. Temporal fidelity was quantified as the lag of the peak cross-correlation between each reconstruction’s LV-area curve and the binned reference across SAX slices (three per patient: apical, basal, and mid). Slices with peak cross-correlation below r = 0.5 were excluded.

**FIGURE 6:**
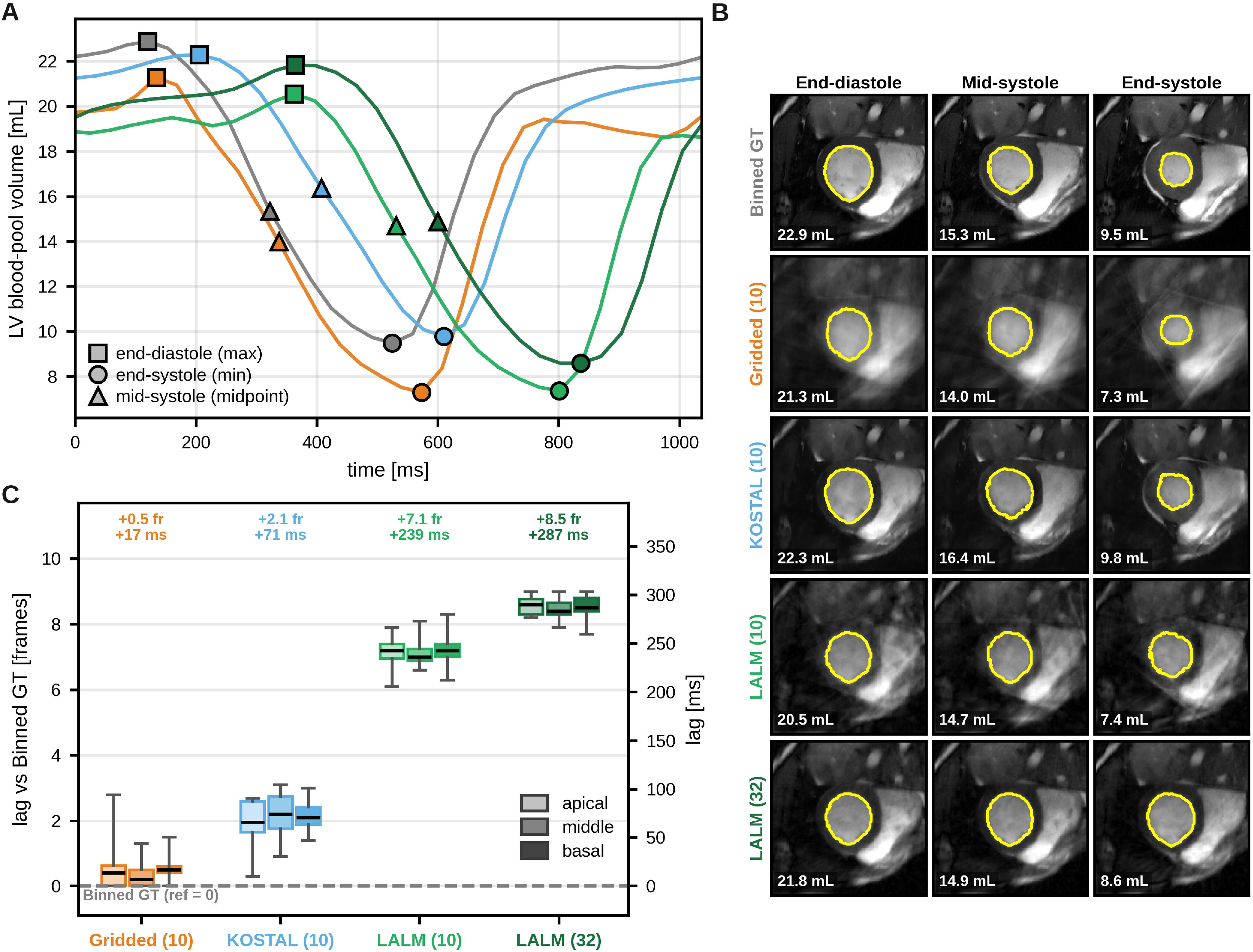
Real-time left-ventricular (LV) function in the prospective cohort. Each method is delayed by the time required to produce its frame, which is the measured reconstruction time rounded up to an integer number of frames. The traces and images therefore report the delay an operator would see at the scanner rather than an idealized instantaneous reconstruction. One frame is 34 ms. **(A)** LV blood-pool volume over one cardiac cycle for a basal SAX slice of one patient, from the binned reference (Binned GT, gray) and the four real-time reconstructions (methods as in Figure 4). Squares mark the end-diastolic maximum, triangles the mid-systolic midpoint, and circles the end-systolic minimum. **(B)** LV endocardial segmentation of that slice at those three instants, one row per reconstruction, each read at the frame that the method had on screen at that moment. **(C)** Cross-correlation lag of the LV-area curve relative to the binned reference from 23 patients (three slices per patient), separated by reconstruction and by anatomical level, the latter encoded as a lightness ramp of the method color (apical lightest, basal darkest). Slices whose cross-correlation peak fell below r = 0.5 were excluded. Boxes span the first to third quartile with the median marked and whiskers at 5^th^ and 95^th^ percentiles. The right-hand axis restates the frame axis in milliseconds, and the dashed line at zero is the binned reference.

For the comparison against the clinical cine (Figure 7, Figure S4), heartbeats were detected in the free-breathing acquisition, with the first and last discarded. A representative single-slice example is shown in Figure 7A-B, where all beats were ranked by stroke volume. The five central-ranking beats were plotted alongside the patient’s corresponding cine curve, with time scaled to a common cardiac-phase time axis to enable accurate volume comparisons. In Figure 7C, each real-time volume represents the median across all detected beats for that slice, with error bars indicating the beat-to-beat standard deviation. Each patient contributed three SAX slices, one basal, one mid-ventricular, and one apical (69 slices from 23 patients). Slices were manually chosen by verifying that the left-ventricular segmentation was correct in both reconstructions at end-diastole and end-systole, a criterion independent of the agreement being measured. Agreement was quantified by the squared Pearson correlation coefficient, the intraclass correlation coefficient ICC(A,1), and Bland-Altman bias (KOSTAL minus cine), with 95% limits of agreement defined as the bias plus and minus 1.96 standard deviations.

**FIGURE 7:**
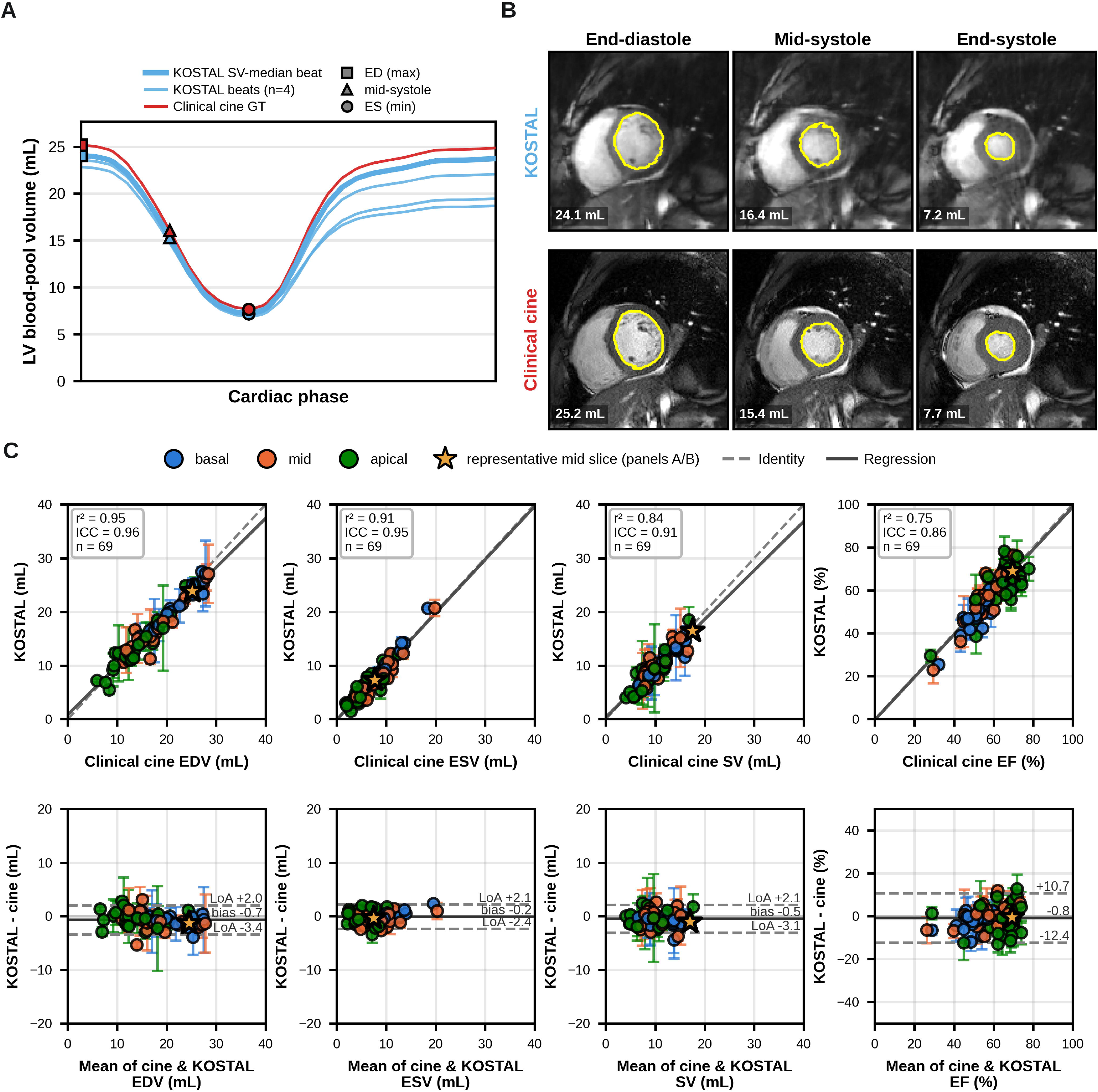
LV function measured on KOSTAL against the clinical breath-held, retrospectively gated cine. **(A)** LV blood-pool volume over one cardiac cycle for a mid-ventricular SAX slice. Heartbeats detected in the free-breathing real-time acquisition were ranked by stroke volume; the bold trace is the stroke-volume median and corresponds to the beat imaged in (B). The clinical cine cycle is overlaid in red, and both were time-warped onto a common cardiac-phase axis so that volume, not timing, is compared. Squares mark end-diastole, triangles mid-systole, and circles end-systole. **(B)** The segmented LV blood pool of that slice at those three instants, for the real-time reconstruction (top) and the clinical cine (bottom). **(C)** Agreement across the cohort, over 69 SAX slices from 23 patients (three per patient). Top row: correlation of the real-time against the clinical-cine end-diastolic volume (EDV), end-systolic volume (ESV), stroke volume (SV) and ejection fraction (EF), with the identity line dashed and the least-squares fit solid. Panels are annotated with the squared Pearson correlation coefficient, the intraclass correlation coefficient (ICC), and n. Bottom row: the corresponding Bland-Altman plots, in which the solid line is the bias and the dashed lines (LoA) the 95% limits of agreement (bias plus and minus 1.96 SD). Each point is one slice, colored by anatomical level; the real-time value is the median over all detected heartbeats of that slice and the error bar is the beat-to-beat SD, whereas the gated cine is a single averaged cycle and carries no spread. The starred point is the slice shown in (A) and (B). Slices were retained only where the LV segmentation was correct in both reconstructions at end-diastole and end-systole, a criterion independent of the agreement being measured.

### Statistical Analysis

All comparisons were paired: each held-out test patient, plane-transition pair, or prospective patient was reconstructed by every method being compared. Because the sample sizes are modest (n = 20 to 33) and the metrics are bounded (SSIM), strictly positive and right-skewed (MSE, recovery time), or expressed as a ratio to a per-patient reference (CNR, ES), differences between reconstructions were assessed with the two-sided Wilcoxon signed-rank test, and all results are summarized as medians with interquartile ranges. Every comparison reported is between KOSTAL and another reconstruction, or between two KOSTAL configurations; comparison methods were not tested against one another. For panels comparing three or more reconstructions, a Friedman test was run first and the pairwise comparisons treated as post hoc. Multiplicity was controlled by the Holm-Bonferroni procedure within each family of comparisons, a family being one panel, one metric, and one cohort; reported p-values are Holm-adjusted. Agreement between the real-time reconstruction and the clinical cine was quantified by the squared Pearson correlation coefficient, the intraclass correlation coefficient ICC(A,1), and Bland-Altman bias with 95% limits of agreement. Statistical significance was set at a two-sided alpha of 0.05. Analyses were performed in Python with SciPy 1.17.0.

## Results

### Model Development

#### KOSTAL’s dual-domain recurrent architecture delivers high coil-combined image quality reconstruction

On the 33 held-out OCMR test series, reconstruction with 10 radial spokes per frame using the k-space recurrent module alone reconstructed the SOS coil-combined image with a median structural similarity index (SSIM) of 0.891 (0.866-0.911) and median mean-square error (MSE) of 0.467 (0.339-0.723) x 10^-3^. This was significantly higher than the 10-spoke gridded input – SSIM: 0.622 (0.574-0.661 x 10^-3^, p<0.001) and MSE: 4.524 (3.604-5.157 x 10^-3^, p<0.001) (Figure 2A, Table 1). The performance matched gridded reconstructions formed from substantially more radial data. The median SSIM matched static gridding of approximately 63 spokes or dynamic gridding of approximately 81 spokes. This suggests k-space KOSTAL processing can offset an undersampling factor of 6.3.

Addition of the image-space module after the k-space module significantly raised median SSIM to 0.907 (0.889-0.927, p<0.001) and lowered MSE to 0.454 (0.325-0.687, p<0.001) x 10^-3^. This is equivalent to approximately 71 and 114 spokes of static and dynamic gridding by SSIM, and 76 and 235 spokes by MSE. Static gridding increases SSIM monotonically toward the 1000-spoke reference; MSE follows the same trend but inverted, with higher quality corresponding to lower MSE values. However, SSIM for dynamic gridding increases with spoke count and then plateaus. This is due to temporal blurring from spokes accrued across successive cardiac frames degrading image quality.

#### Recurrent unit architecture has a small but consistent effect on image quality

The choice of recurrent architecture had a small but significant influence on reconstruction quality (Figure 2C). For the k-space module, all subtypes performed well (SSIM > 0.83, MSE < 1.2 x 10^-3^), with the GRU performing highest on SSIM (0.856 versus 0.835 for RNN and 0.836 for LSTM, both p<0.001) and lowest on MSE (0.797 versus 1.136 and 1.033 x 10^-3^, both p<0.001).

A recurrent module in image space improved IQ but values were significantly lower (SSIM 0.646-0.650, p<0.001, MSE 4.30-4.52 x 10^-3^, p<0.001) than the k-space module. Further, the same relative performance differences based on architecture were observed. When the recurrent image-space module was added on to the k-space GRU, the GRU again yielded the highest SSIM (0.909 versus 0.902 for RNN and 0.900 for LSTM, both p<0.001) and lowest MSE (0.381 versus 0.436 and 0.444 x 10^-3^, both p<0.001).

Of note, the image-domain KOSTAL module was trained with k-space processing but applied to the 10-spoke gridded data (Fig 2A). Despite this, performance was similar to that achieved by the image-domain GRU trained directly on the 10-spoke gridded data (Fig 2C). This suggests that the limited performance of the image-domain KOSTAL network is not due to a mismatch between the training data (fuller k-space) and testing environment but rather appears to be a limitation of the image-space processing approach itself.

## Testing

### Coil-wise and coil-combined performance are comparable

Table 1 reports per-coil metrics alongside the SOS coil-combined analysis. The ordering of the reconstructions was preserved under both combination strategies: the coil-wise average gave a median SSIM of 0.921 (0.876-0.943) for the k-space output and 0.945 (0.915-0.960) for the combined output, with MSE of 0.106 (0.064-0.168) and 0.081 (0.055-0.142) x 10^-3^. The per-coil reconstructions were at least as accurate as the SOS images formed from them. Both comparisons remained significant under the coil-wise average, as they were under the SOS combination: the k-space output versus the 10-spoke gridded input, and the combined output versus the k-space output (all p<0.001).

### Recurrent state enables rapid recovery of image quality after through-plane transitions

KOSTAL’s recurrent state enables rapid recovery of image quality following a sudden, drastic change in imaging data (Figure 3). Under the SSIM criterion, KOSTAL recovered image quality in 75 (66-92) ms. This was significantly faster than equivalent IQ gridded approaches (111 (94-125) ms for 75-spoke gridding and 303 (252-337) ms for 200-spoke gridding; both p<0.001). Analysis using MSE, identifies recovery as occurring in 77 (61-94) for KOSTAL, 140 (125-154) for 75-spoke gridding, and 388 (353-431) ms for 200-spoke gridding (both p<0.001).

## Validation

### KOSTAL yields the highest image quality across the prospective cohort

Representative reconstructions from KOSTAL and the comparison methods are shown in Figure 4A. Without any retraining or fine-tuning, KOSTAL robustly reconstructed all acquired anatomies, including the multi-slice SAX stack and the two-, three-, and four-chamber LAX views. Respiratory-and cardiac-binned reconstruction of the same golden-angle data provided a fully sampled internal reference from the same acquisition. Relative to this binned reference, KOSTAL achieved the best agreement of all methods across SSIM and MSE (Fig. 4B), exceeding every comparison method on both metrics and in both orientations (all p<0.05).

KOSTAL achieved the highest CNR in the LAX views (all p<0.01) and higher edge sharpness (ES) than the iterative methods in both orientations (all p<0.001) (Figure 5B). Expressed as a percentage of each patient’s binned reference, KOSTAL reached a median CNR of 84% SAX and 87% LAX. Only the 32-spoke iterative reconstruction was comparable, marginally exceeding KOSTAL SAX (86%, p=0.87) while falling below in LAX (78%, p<0.001). MDCNN attained higher ES in the SAX views (109%, p<0.001) and comparable ES in the LAX views (107%, p=0.29) but exhibited clear image-quality artifacts.

### KOSTAL inference adds only a small computational cost and is markedly faster than iterative reconstruction

KOSTAL’s dual-domain recurrent GRU reconstruction adds only a modest computational cost to the gridding pipeline (Fig. 4C-D). Because the recurrent modules operate per-coil and are readily parallelized on a graphics processing unit (GPU), total inference time is fast (<34 ms per frame) and only marginally slower (18ms) than gridding alone, despite KOSTAL’s larger computational burden (∼2x compared to MDCNN). Further, KOSTAL’s design allows it to process any number of coils. On the other hand, processing time increases with LALM, and the number of coils processed in a subset is determined by the hardware available. For MDCNN, a different number of coils would require retraining. With an 8-coil input, both KOSTAL and MDCNN processing times remained below that of 1-frame display lag (reconstruction time shorter than the 10-radial-spoke acquisition time). The LALM approaches have display lags of 8-9 frames when processing 8 coils.

### KOSTAL’s 10-spoke reconstruction tracks left-ventricular blood-pool volume with small errors and lower latency than wider-footprint gridded reconstructions

KOSTAL tracked the left-ventricular blood-pool volume curve of the binned reference with a median lag of 2.1 frames (71 ms), against 0.5 frames (17 ms) for 10-spoke gridding and 7.1 and 8.5 frames (239 and 287 ms) for the 10-and 32-spoke iterative reconstructions (Figure 6). KOSTAL therefore incurred only a 1-to 2-frame lag relative to 10-spoke gridding while providing substantially higher image quality. The same behavior held at basal and apical levels (Figure S3).

KOSTAL produced slice volumes that consistently matched the cine reference (Figure 7A-B; Figure S4). Over 69 SAX slices from 23 patients, correlation and Bland-Altman analyses showed close agreement for end-diastolic and end-systolic volume (EDV, ESV) and weaker agreement for the derived stroke volume (SV) and ejection fraction (EF) (Figure 7C). Biases were small:-0.7 mL for EDV (95% limits of agreement-3.4 to +2.0 mL),-0.2 mL for ESV (-2.4 to +2.1 mL),-0.5 mL for SV (-3.1 to +2.1 mL) and-0.8 percentage points for EF (-12.4 to +10.7).

## Discussion

A dual-domain recurrent convGRU-based algorithm (KOSTAL) can reconstruct 10-spoke golden-angle radial CMR at an image quality conventionally achieved by gridding 7-11x more spokes, in only 34 ms per frame. Ablation studies indicate that most of the gain comes from the k-space processing module. Specifically, a recurrent module operating on gridded k-space raised SSIM from 0.622 to 0.891, whereas the same module operating in the image domain only raised image quality to at most 0.650. This difference far exceeded that observed across different recurrent network architectures. This supports the premise that exploiting temporal aspects of a golden-angle acquisition in the sampling domain itself, where a gate conditioned on the binary mask can decide, location by location, whether to retain the accumulated sample or take the new one, is beneficial. This agrees with prior work that leverages view-sharing^29,30^. However, while those algorithms define data-merging rules, KOSTAL learns them during the training process.

This k-space-centric approach also distinguishes KOSTAL from existing recurrent reconstructions. Jaubert et al. placed convLSTM blocks inside an image-domain U-Net and achieved comparable inference time. However, they reported that image quality takes several frames to stabilize and that abrupt orientation changes produce transient morphing artifacts lasting approximately half a second^40^. Other deep learning approaches similarly report suboptimal image quality during interactive scan-plane changes^14,16^. These prior results motivated our direct evaluation of this potential failure mode in our plane-transition experiments. Our learned gating recovers from a plane transition in 75 ms, faster than 75-spoke gridding while matching performance at steady state. Other approaches explore temporal information but differ in significant technical aspects. Qin et al. exploit temporal redundancy bidirectionally but this precludes causal, real-time imaging deployment^41^. Further, recurrent variational networks and LORAKI recur over unrolled optimization steps rather than over acquisition time, and the convLSTM approach of Zhao et al requires a pre-operative reference image^42–44^. KOSTAL’s recurrent state in the k-space domain is the accumulated k-space itself, and it does not require prior scan data.

These advantages were further demonstrated in the prospective validation results. Without retraining, KOSTAL yielded the best agreement with the binned reference, the highest LAX CNR, and an LV-area lag of 74 ms against 243 and 287 ms for iterative reconstructions. The performance falls comfortably inside the 200 ms latency budget of iCMR^10,11^.

Several limitations qualify these findings. The network was trained by resampling OCMR Cartesian data, with radial sampling simulated retrospectively from breath-held Cartesian bSSFP. The fact that this simulated training transferred well to prospective free-breathing data without fine-tuning is encouraging, but it does not establish that training on prospective data would not perform better^45^. Second, the prospective cohort lacks fully sampled, ECG-gated golden-angle radial reference data. As a result, the binned XD-GRASP approach was used as a reference^38^. Another limitation is that the validation cohort is relatively small and from a single center and single vendor. However, these promising results motivate multi-vendor and multi-site evaluation, inline deployment, and evaluation during interventional guidance. Further, arrhythmias were not directly evaluated. A related question is whether a network trained largely on structurally typical hearts might bias reconstruction of atypical anatomy toward a more normal appearance. Because KOSTAL’s recurrent state is the accumulated k-space of the acquisition itself and supervision is applied primarily in k-space, we expect the network to learn generic sampling and view-combination operations rather than anatomic priors. The successful use of the trained network to the independent pediatric RCHSD cohort without fine-tuning is consistent with this. However, this has not been tested directly, and dedicated evaluation in cohorts with abnormal anatomy, including single-ventricle physiology, is warranted. Finally, reconstruction was done offline. Therefore, latency was modeled based on measured component times rather than observed in a live inline loop. The times and ability to process coils in parallel depend on the hardware utilized. We expect this could significantly improve with use of even higher-end hardware and advances in compute in the near future.

## Conclusions

A recurrent-based, dual-domain neural network operating directly on k-space recasts view-sharing as a learned, mask-conditioned operation, and delivers 10-spoke real-time cardiac reconstruction at the quality of a much larger acquisition, with a latency compatible with interventional guidance.

## Data Availability

Data and software will be made available upon reasonable request.

## Software code

Upon publication, inference software will be made available via Github.

