## Supplemental Figures 1-4 for "K-space Optimized Spatial Temporal Architecture for Low Latency in Interventional Cardiac MRI"

### Contents

**Supporting Figure S1:** *Ablation of the five loss terms in KOSTAL architecture.*

**Supporting Figure S2:** *Cumulative energy versus coil-compression depth for SVD and ROVir for the prospective validation cohort.*

**Supporting Figure S3:** *Basal and apical companions to Figure 6, from two further patients.*

**Supporting Figure S4:** *LV blood-pool volume-time curves for the prospective validation cohort.*

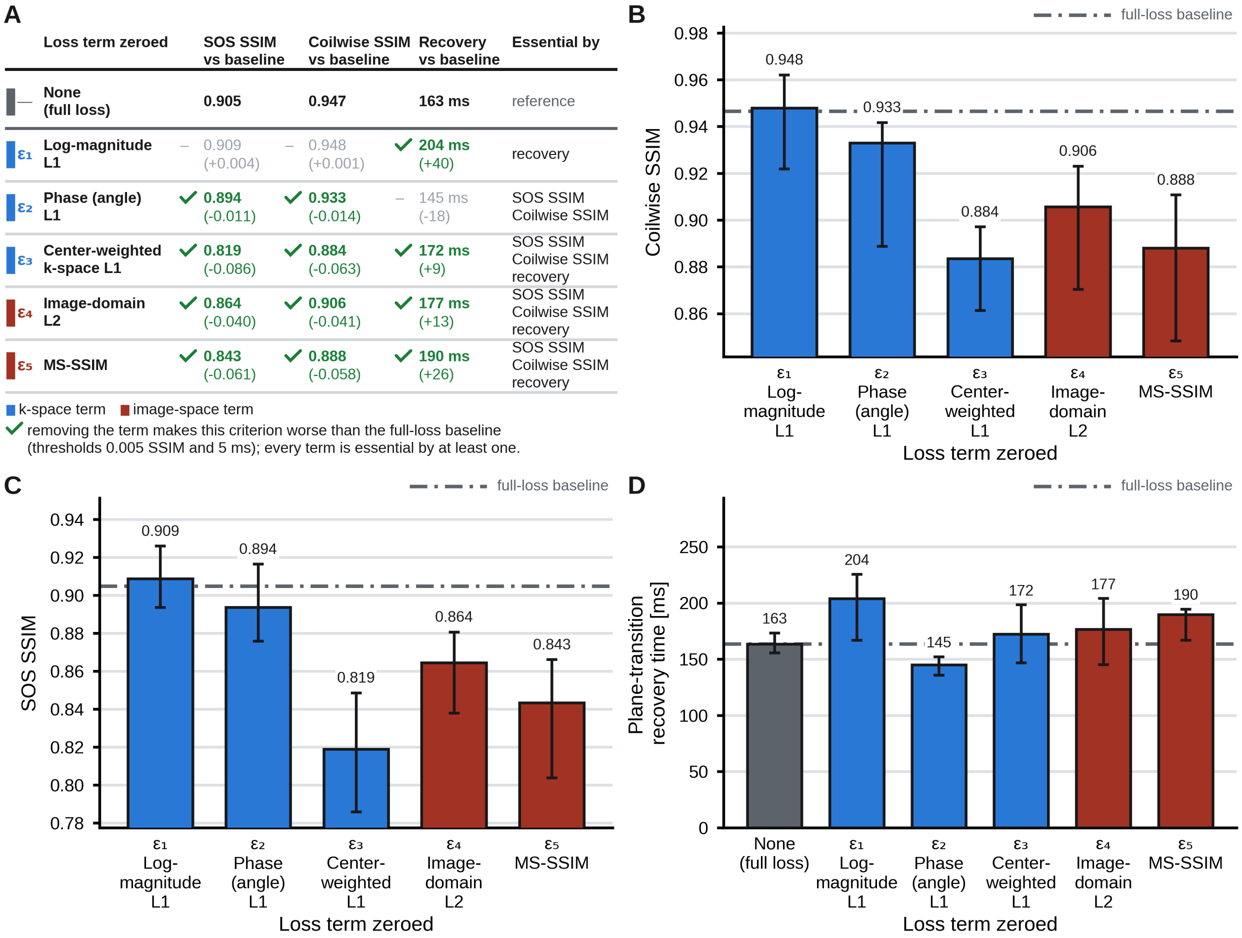

| | Loss term zeroed | SOS SSIM vs baseline | Coilwise SSIM vs baseline | Recovery vs baseline | Essential by |
|---|---|---|---|---|---|
| — | None (full loss) | 0.905 | 0.947 | 163 ms | reference |
| ε1 | Log-magnitude L1 | – 0.909 (+0.004) | – 0.948 (+0.001) | ✓ 204 ms (+40) | recovery |
| ε2 | Phase (angle) L1 | ✓ 0.894 (-0.011) | ✓ 0.933 (-0.014) | – 145 ms (-18) | SOS SSIM Coilwise SSIM |
| ε3 | Center-weighted k-space L1 | ✓ 0.819 (-0.086) | ✓ 0.884 (-0.063) | ✓ 172 ms (+9) | SOS SSIM Coilwise SSIM recovery |
| ε4 | Image-domain L2 | ✓ 0.864 (-0.040) | ✓ 0.906 (-0.041) | ✓ 177 ms (+13) | SOS SSIM Coilwise SSIM recovery |
| ε5 | MS-SSIM | ✓ 0.843 (-0.061) | ✓ 0.888 (-0.058) | ✓ 190 ms (+26) | SOS SSIM Coilwise SSIM recovery |



**SUPPORTING FIGURE S1:** *Ablation of the five loss terms in KOSTAL architecture.*

Each ablated model was trained from scratch with one loss weight set to zero and evaluated on the 33 held-out test patients and the 33 held-out plane-transition pairs against the full-loss baseline. **(A)** Summary table giving, for each zeroed term, the median SOS SSIM, the median coil-wise SSIM, and the median plane-transition recovery time, with the change relative to baseline in parentheses. A term was judged essential on a criterion when its removal worsened that criterion by more than 0.005 SSIM or 5 ms. Blue marks a k-space term and red an image-space term, as in Figure 1B. **(B, C)** Median coil-wise (B) and SOS (C) SSIM of the combined k-space plus image-space output against the 1000-spoke reference, over the central 50% cardiac ROI; bars are medians, whiskers are the first to third quartile, and the dash-dotted line is the full-loss baseline. **(D)** Median time to recover 75% of the plateau SOS SSIM after a change of imaging plane. Whiskers are the first to third quartiles, and the baseline is drawn both as a gray bar and as the dash-dotted line.

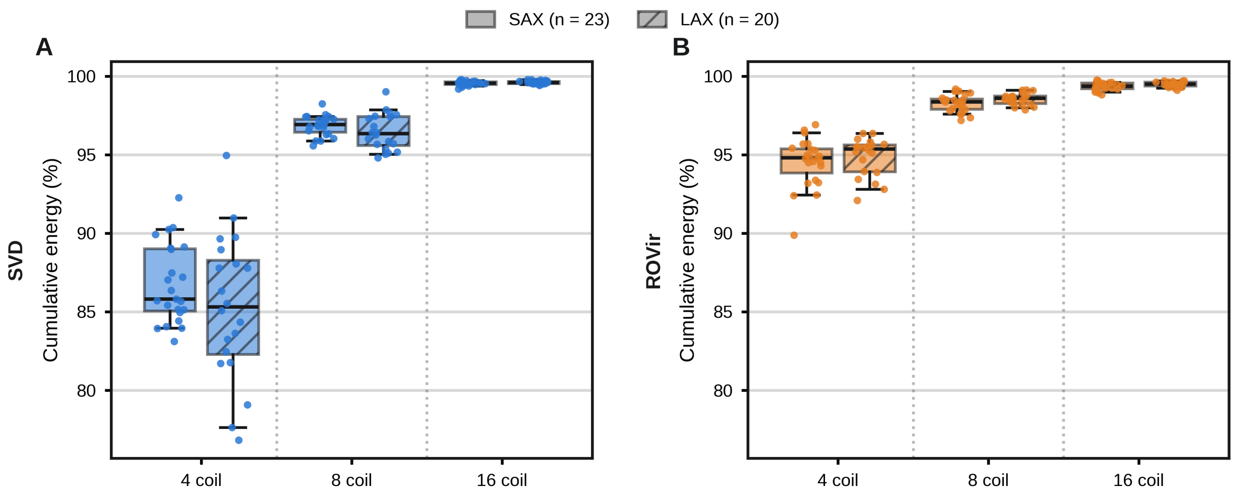


**SUPPORTING FIGURE S2:** *Cumulative energy versus coil-compression depth for SVD and ROVir for the prospective validation cohort.*

**(A, B)** Each panel plots the cumulative fraction of the compression spectrum retained at three fixed coil budgets (4, 8 and 16 virtual coils); dotted lines separate the budgets, and within each budget the SAX and LAX acquisitions are drawn side by side. Color identifies the compression method (SVD left, ROVir right). Each point represents one patient, taken as the median across that patient's slices (SAX, n = 23; LAX, n = 20); boxes span the first to third quartiles, with the median marked, and whiskers the 5$^{th}$ to 95$^{th}$ percentiles.

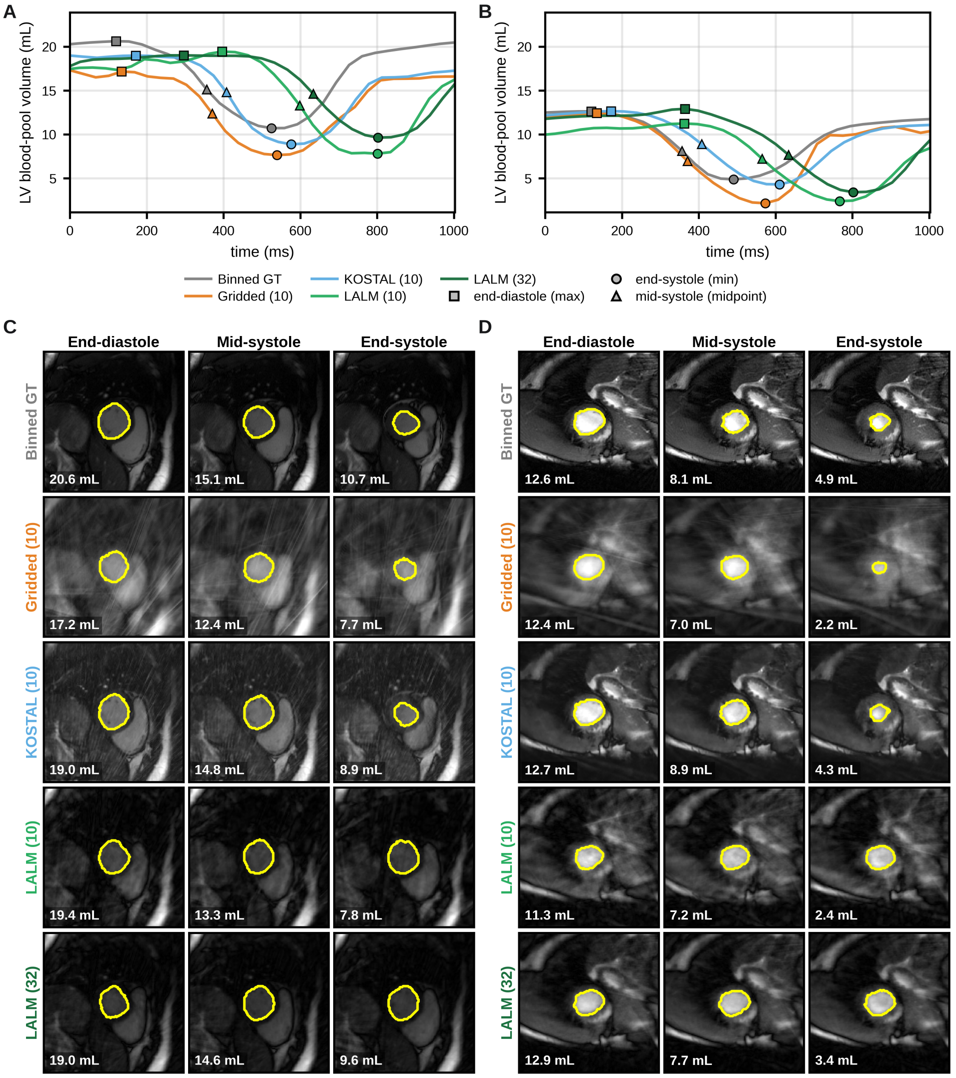


**SUPPORTING FIGURE S3:** *Basal and apical companions to Figure 6, from two further patients.*

As in Figure 6, every reconstruction is displayed through its own reconstruction latency. **(A, B)** LV blood-pool volume against time for a basal (A) and an apical (B) SAX slice, from the binned reference (Binned GT) and the four real-time reconstructions. Squares mark the end-diastolic maximum, triangles the mid-systolic midpoint and circles the end-systolic minimum. The two panels share one volume axis, so basal and apical volumes are directly comparable. **(C, D)** LV endocardial segmentations of the basal (C) and apical (D) slices at those three instants, one row per reconstruction, each read at the frame that the method had on screen at that moment.

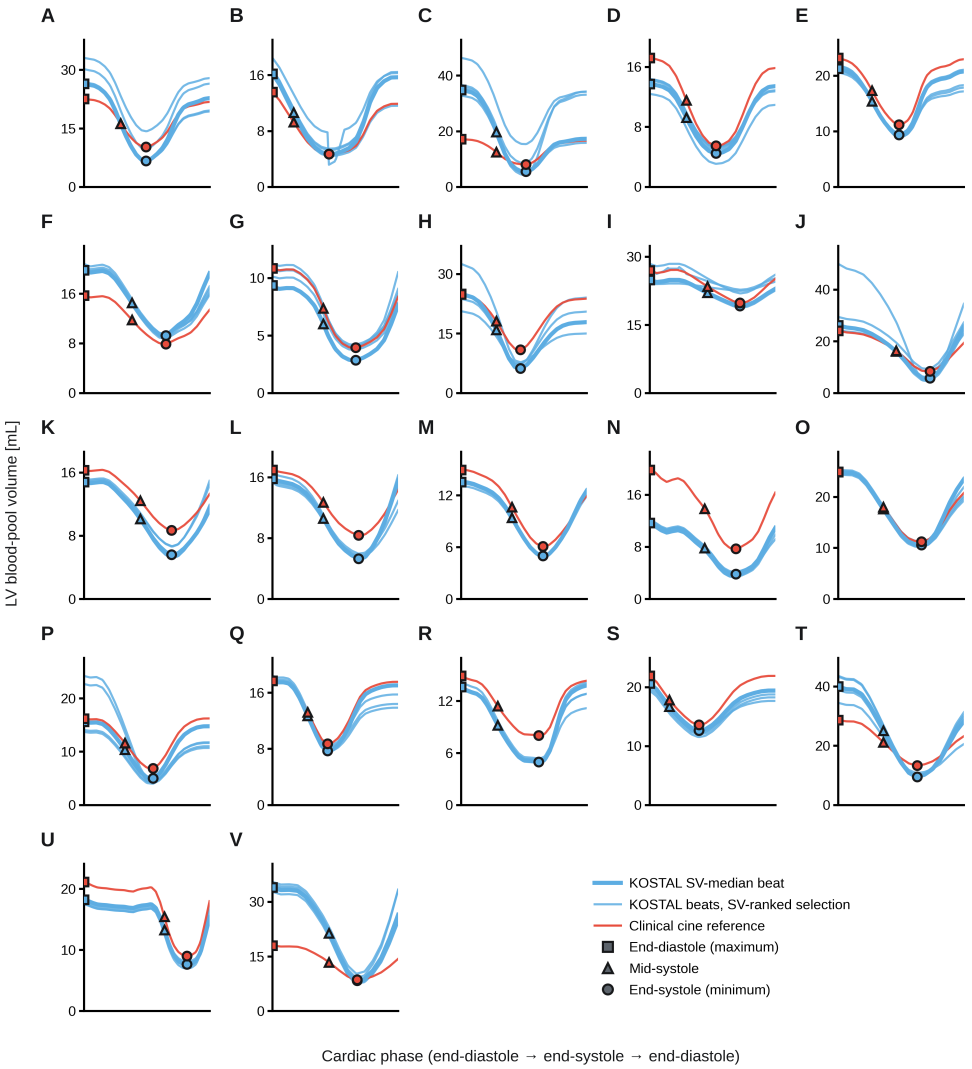


**SUPPORTING FIGURE S4:** *LV blood-pool volume-time curves for the prospective validation cohort.*

**(A-V)** One panel per patient: the twenty-third patient is shown in full in Figure 7A-B and is not repeated here. For each patient, the mid-ventricular SAX slice is shown. Heartbeats detected in the free-breathing KOSTAL acquisition were ranked by stroke volume; the bold blue trace is the stroke-volume-median beat of that selection. The paired clinical breath-held, retrospectively gated cine cycle is overlaid in red. All curves were time-warped onto a common cardiac-phase axis running from end-diastole through end-systole back to end-diastole, so that volume and not timing is compared; squares mark end-diastole, triangles mid-systole and circles end-systole. Each panel carries its own volume axis, scaled to that patient's volumes, so trace heights are not comparable between panels.
